# Can machine learning improve risk prediction of incident hypertension? An internal method comparison and external validation of the Framingham risk model using HUNT Study data

**DOI:** 10.1101/2022.11.02.22281859

**Authors:** Filip Emil Schjerven, Emma Ingeström, Frank Lindseth, Ingelin Steinsland

**Affiliations:** Department of Computer Science, Norwegian University of Science and Technology, Trondheim, Norway; Department of Circulation and Medical Imaging, Norwegian University of Science and Technology, Trondheim, Norway; Department of Mathematical Sciences, Norwegian University of Science and Technology, Trondheim, Norway

## Abstract

A recent meta-review on hypertension risk models detailed that the differences in data and study-setup have a large influence on performance, meaning model comparisons should be performed using the same study data. We compared five different machine learning algorithms and the externally developed Framingham risk model in predicting risk of incident hypertension using data from the Trøndelag Health Study. The dataset yielded *n* = 23722 individuals with *p* = 17 features recorded at baseline before follow-up 11 years later. Individuals were without hypertension, diabetes, or history of CVD at baseline. Features included clinical measurements, serum markers, and questionnaire-based information on health and lifestyle. The included modelling algorithms varied in complexity from simpler linear predictors like logistic regression to the eXtreme Gradient Boosting algorithm. The other algorithms were Random Forest, Support Vector Machines, K-Nearest Neighbor. After selecting hyperparameters using cross-validation on a training set, we evaluated the models’ performance on discrimination, calibration, and clinical usefulness on a separate testing set using bootstrapping. Although the machine learning models displayed the best performance measures on average, the improvement from a logistic regression model fitted with elastic regularization was small. The externally developed Framingham risk model performed well on discrimination, but severely overestimated risk of incident hypertension on our data. After a simple recalibration, the Framingham risk model performed as well or even better than some of the newly developed models on all measures. Using the available data, this indicates that low-complexity models may suffice for long-term risk modelling. However, more studies are needed to assess potential benefits of a more diverse feature-set. This study marks the first attempt at applying machine learning methods and evaluating their performance on discrimination, calibration, and clinical usefulness within the same study on hypertension risk modelling.

**Author summary:** Hypertension, the state of persistent high blood pressure, is a largely symptom-free medical condition affecting millions of individuals worldwide, a number that is expected to rise in the coming years. While consequences of unchecked hypertension are severe, life-style modifications have been proven to be effective in prevention and treatment of hypertension. A possible tool for identifying individuals at risk of developing hypertension has been the creation of hypertension risk scores, which calculate a probability of incident hypertension sometime in the future. We compared applying machine learning as opposed to more traditional tools for constructing risk models on a large Norwegian cohort, measuring performance by model validity and clinical usefulness. Using easily obtainable clinical information and blood biomarkers as inputs, we found no clear advantage in performance using the machine learning models. Only a few of our included inputs, namely systolic and diastolic blood pressure, age, and BMI were found to be important for accurate prediction. This suggest more diverse information on individuals, like genetic, socio-economic, or dietary information, may be necessary for machine learning to excel over more established methods. A risk model developed using an American cohort, the Framingham risk model, performed well on our data after recalibration. Our study provides new insights into machine learning may be used to enhance hypertension risk prediction.

## Introduction

Individuals with persistently high levels of blood pressure are said to have hypertension. It is a mostly symptom-free medical condition, but it increases the risk of more severe diseases and premature death if left untreated (1). The number of hypertensive individuals is estimated to be over 1 billion worldwide, and suboptimal blood pressure accounts for around 10 % of the world’s overall health expenditures (2,3). It is well-established that the risks related to hypertension can be effectively reduced through lifestyle modifications and medications (1). Hence, early identification of otherwise healthy individuals at risk of hypertension has become a multidisciplinary research effort including the analysis and subsequent development of hypertension risk models.

Since the publication of the Framingham risk model for incident hypertension in 2009, the number of risk models has increased substantially and the topic has been reviewed multiple times (4–6). In a recent review, 52 studies and 117 risk models were identified, of which most models were developed using established statistical methods (7). Simultaneously, machine learning has emerged as a common alternative for constructing risk models. By leveraging the ability to learn more complex patterns from the data with less human intervention, machine learning has been emphasized as having the potential to construct models excelling those using less-complex methods. Machine learning has been shown to improve risk prediction for cardiovascular disease (8). Many speculate whether machine learning and other artificial intelligence (AI) methods can contribute to transforming the health-sciences (9) (10).

However, this potential has been challenged in studies reviewing risk prediction models for other diseases. In these cases, applying machine learning to a problem was not synonymous with improved performance compared to more established statistical methods (11–13). When not restricting to a specific disease, a systematic review by Christodoulou et al. found that logistic regression performed just as well as machine learning for clinical prediction models when limiting themselves to studies with low risk of bias (14).

Considering the topic of hypertension risk models specifically, a recent systematic review identified and summarized the previous work (7). In that review, it was found that a large variation reported in discrimination performance was unrelated to the type of modelling algorithms being used. In other words, using different datasets and study setups had considerable influence on the reported performance. This implies that to confidently determine a performance advantage of one method over the other, it would be necessary to compare methods within the same study and data, keeping everything fixed except the methods under investigation. Thus, it is difficult to determine the cause of any apparent performance benefit that has been reported for one method over others in the existing literature.

For a prediction model to be useful to health practitioners in the real-life clinical setting, it must demonstrate acceptable performance during evaluation. A model’s performance is often captured by its discriminative performance, as well as the calibration of its predictions. Furthermore, clinical usefulness has been emphasized to demonstrate that the risk models provide a clear benefit over alternatives in decision-making (15–17). In the literature on hypertension risk models, discrimination is the most frequently reported performance indicator. Although calibration is often reported for risk models developed using more established methods, few studies using machine learning have reported model calibration performance. Lastly, few studies have explored clinical usefulness for hypertension risk models using decision curve analysis like Net Benefit (7,15,17).

Machine learning have been used to develop hypertension risk models before, but few studies have compared their performance with low-complexity models like logistic or Cox regression (18–20). Among the studies who have, machine learning has been found to have better discrimination in some studies and worse in others (11,21–24). Calibration was assessed in only two studies applying machine learning models, and then solely by reporting a summary measure (22,25). Niu et al. evaluated machine learning models by their net benefit as well as discrimination, and displayed a large advantage of using machine learning models on a rural Chinese cohort (24).

Considering the many existing risk models, a new model should be a valuable contribution to the literature. External validation, as opposed to creating new risk models, has been emphasized as an equally valuable contribution to the literature and a necessity for transitioning a risk model to clinical practice. (26–28). Not only may it provide information on how well the existing model generalizes to new, unseen data, but it also serves as a benchmark for the development of any new proposed model.

In this work, we aim to investigate the potential performance benefit of using machine learning for predicting hypertension by conducting a thorough method comparison. Our primary aim was to assess whether more complex modelling methods provide a clear performance benefit, in terms of discrimination, calibration, and clinical usefulness, over more established statistical methods when developed under equal settings using the same data. To assess the need for a new risk model and provide benchmarks for our comparison, we externally validated the Framingham risk model (4).

## Results

### Developed models

Summary statistics for our study data are provided in Table in S2 Table. There were significant differences in all features when stratified on outcome status. Between the training and testing set, BMI and Physical Activity had small, but significant differences between means or proportions between the training and testing sets. All other variables had no significant difference, see Table in S6 Table. The outcome rate of the full, training and testing set was 24.65%, 24.36% and 25.3%, respectively. Missing entries in the features were relatively low for almost all features. Missingness exceeded 10 % for ‘*Family history of hypertension’* and 1% for ‘*Family history of CVD’*, ‘*Socio-economic status’* and ‘*Physical Activity’*. In total, 4572 individuals missed one entry, 570 missed two, 73 missed three, 9 missed four, and 1 missed five. In sum, 5225 (22%) individuals had at least one missing entry.

Hyperparameters selected from the cross-validation procedure and the results from evaluation on the training set are listed in Table in S7 Table.

Applying the final models upon the testing set, we obtained the bootstrapped results given in Table 1. On average, the eXtreme Gradient Boosting (XGBoost) model performed better on the Area Under the Curve measure (AUC) and Brier score, but with largely overlapping confidence intervals compared to the elastic regression and Support Vector Machine (SVM) models. The Random Forest (RF) model excelled on the Integrated Calibration Index (ICI) and outperformed all other developed models on average.

**Table 1:** Model results achieved on test set.

| Models | AUC (↑) | Brier (↓) | ICI (↓) |
| --- | --- | --- | --- |
| Elastic regression | 0.801 [0.789, 0.813] | 0.148 [0.143, 0.153] | 0.023 [0.015, 0.032] |
| XGBoost | **0.803 [0.791, 0.815]** | **0.147 [0.142, 0.152]** | 0.018 [0.010, 0.026] |
| RF | 0.789 [0.776, 0.802] | 0.152 [0.147, 0.157] | 0.013 [0.006, 0.020] |
| SVM | 0.798 [0.785, 0.810] | 0.150 [0.145, 0.155] | 0.029 [0.021, 0.037] |
| KNN | 0.786 [0.773, 0.799] | 0.153 [0.148, 0.158] | 0.019 [0.011, 0.027] |
| Framingham risk model (Ext.) | 0.794 [0.782, 0.805] | 0.175 [0.170, 0.180] | 0.127 [0.118, 0.136] |
| Framingham risk model, recalibrated (Ext.) | 0.794 [0.782, 0.805] | 0.150 [0.145, 0.154] | **0.011 [0.003, 0.018]** |

Performance obtained applying the fitted models on the test set. Reported as mean and 95% confidence interval after bootstrapping. The symbols (↑) and (↓) signify increasing or decreasing values as improved performance, respectively. Models marked (Ext.) are externally developed models with or without recalibration.

The calibration plots are given in Fig 1 with individual curves and prediction distributions given in S11 Fig. The shape of the curves was similar for all developed models, with most models overestimating risk for predictions above 50%. The RF model obtained a lower ICI as it was overall well-calibrated for predictions below 50%, and far closer to the calibration reference line for most of its predictions compared to the other models.

**Fig 1.**
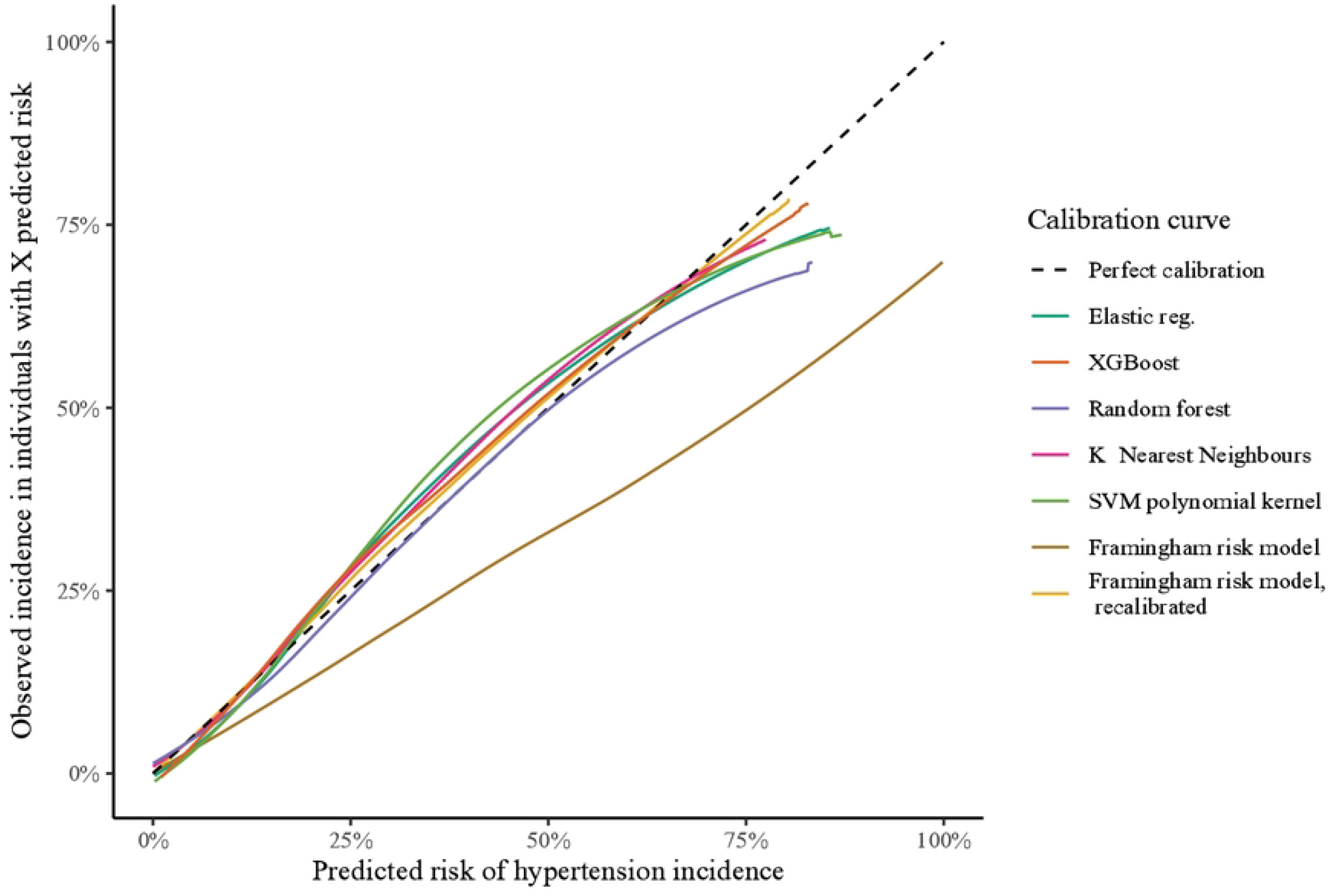
Calibration curves calculated on the test set. Calculated as mean calibration curve after pointwise bootstrapping.

The Net Benefit plot is given in Fig 2 with individual curves given in S12 Fig. Assuming a threshold probability < 50%, all models displayed favorable net benefit compared to the reference options of ‘treat all’, ‘treat none’, or predicting individuals to be ‘Hypertensive’ at follow-up if they were prehypertensive at baseline. Assuming a threshold above 50%, some of the models exhibited negative benefit, i.e., that the cost of erroneous predictions is higher than the benefit. Reviewing net benefit across all thresholds, the elastic regression and XGBoost models yielded the highest, but similar, net benefit.

**Fig 2.**
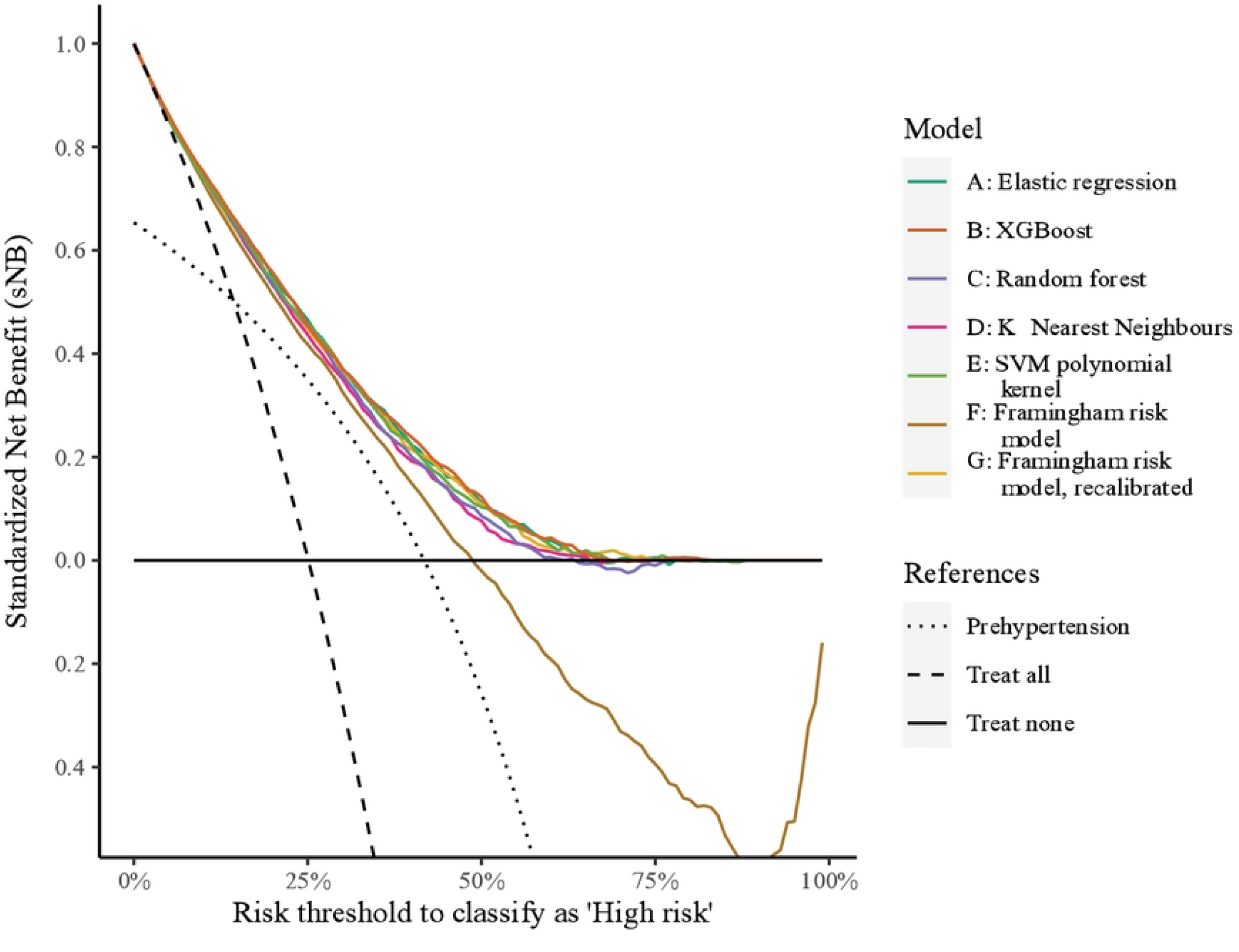
Standardized Net Benefit calculated on the test set. Standardized Net Benefit displays the benefit of applying a model on a population-level and has an upper bound of 1 with no lower bound. A model should be preferred over another if it dominates over all the relevant risk thresholds. See Vickers et al. for further details on interpretation of Net Benefit (72).

Other performance indicators from evaluations on the testing set are reported in Table in S8 Table. In short, XGBoost and elastic regression yielded the best average scores of the developed models.

### External model

Judging by the mean AUC, the Framingham risk model performed comparably to our newly developed models, even outperforming the K-Nearest Neighbor (KNN) and RF models. However, the Framingham risk model had the worst Brier score and calibration compared to the other models. Judging by the calibration plot in Fig 2, the Framingham risk model overestimated risk and increasingly so as the predicted risk became larger.

The Framingham risk model provided favorable Net Benefit compared to the references but was dominated by all other models. Other performance indicators for the Framingham risk model evaluations are listed in Table in S8Table. The average performance was slightly worse when evaluated on the full dataset, but within the 95% confidence interval reported for the testing set, see Table 2. After recalibration of the Framingham risk model using the training set, the recalibrated model achieved a better ICI than all other models on the testing set, see Table 1. The Brier score was comparable, but not superior, to developed models as its discriminatory performance was slightly worse than the other models. See Table in S4 Table for details on the recalibrated model.

**Table 2:** Framingham risk model results achieved on the whole dataset.

| Models | AUC (↑) | Brier Score (↓) | ICI (↓) |
| --- | --- | --- | --- |
| Framingham risk model | 0.787 [0.780, 0.793] | 0.178 [0.175, 0.181] | 0.133 [0.127, 0.138] |

Performance obtained applying the Framingham risk model with adaptations on the whole dataset. Reported as mean and 95% confidence interval after bootstrapping. The symbols (↑) and (↓) signify increasing or decreasing values as improved performance, respectively.

### Sensitivity analysis

The results from our sensitivity analysis using the LASSO regression with increasing penalty are displayed in Fig 3. Most of the coefficients were zeroed out at a low penalty. The order and penalty in which coefficients were zeroed out are given in Table in S9 Table. Diastolic blood pressure, systolic blood pressure, age, and BMI stand out as important features. The AUC performance indicator decreased slowly until these variables were eliminated. However, the Brier and ICI score became notably worse at a lower regularization penalty compared to the AUC score.

**Fig 3.**
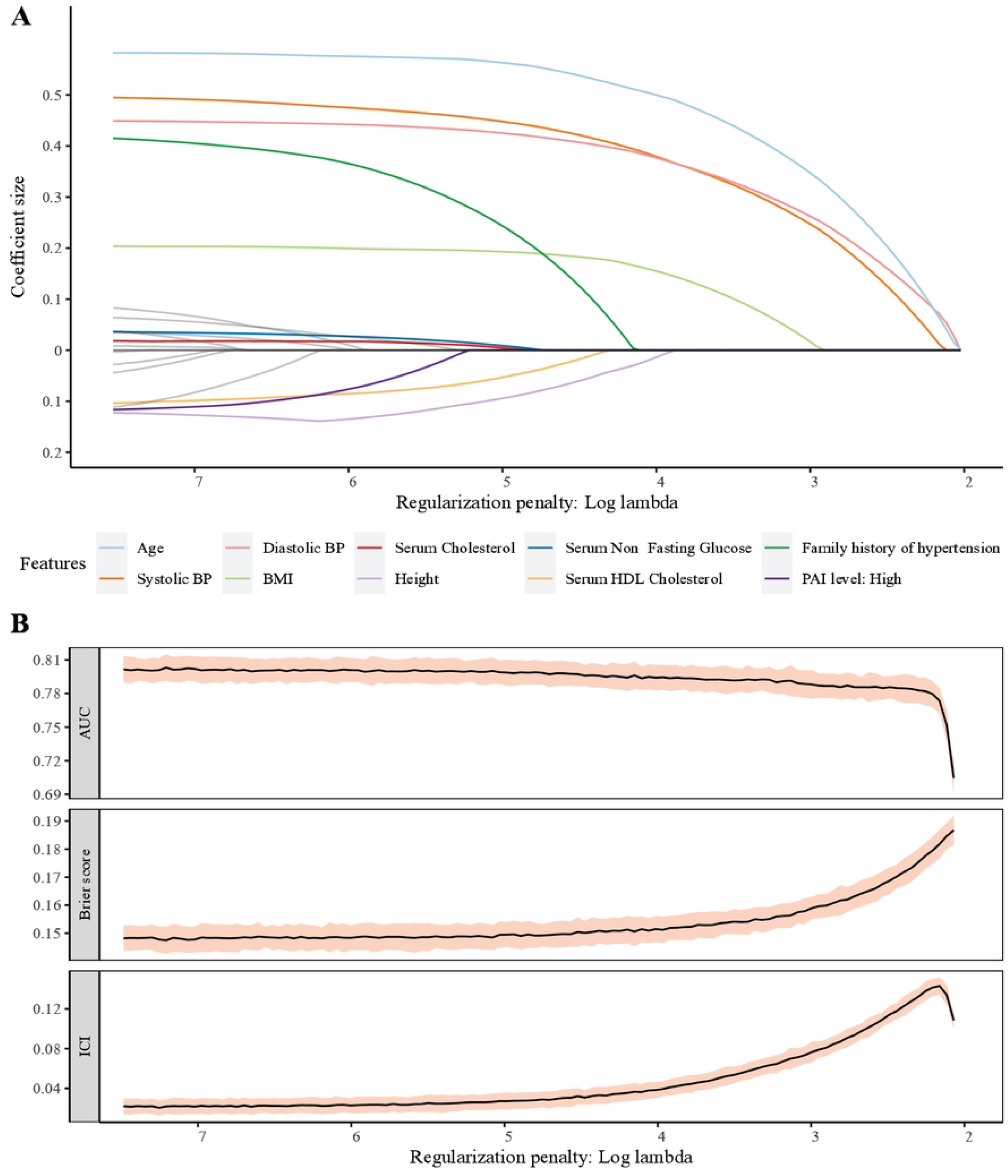
Sensitivity analysis with LASSO regression. (A): Coefficient sizes versus penalty in LASSO regression fitted on the training set. Ten last coefficients to be zeroed out are colored and named. Note, numerical features have been standardized, i.e., coefficient sizes correspond to the increase of one standard deviation of the training set, see Table in S6 Table. (B): AUC, Brier score and ICI calculated on test set using LASSO models with coefficients as in Panel A, with panels corresponding on the X-axis. Black mean line and red 95% confidence interval derived from pointwise bootstrapping. Note, the performance scores as all coefficients are zeroed out have been cropped out as the AUC becomes 0.5, Brier Score 0.1875 and ICI undefined.

To compare the importance of features, we report the normalized importance gain calculated in the XGBoost and RF model from model-fitting on the training set. These are displayed in Fig 4 and reported in detail in Table in S9 Table. In the XGBoost model, the feature importance was concentrated around age, diastolic blood pressure, systolic blood pressure, and BMI, with all others less important. The same top four was reported for the RF model, however, the impact of all numerical measurements were rated higher. BMI was estimated to be less important than age, diastolic blood pressure, and systolic blood pressure, but more important than the others in the LASSO regressions and by the normalized importance gain in XGBoost and RF models.

**Fig 4.**
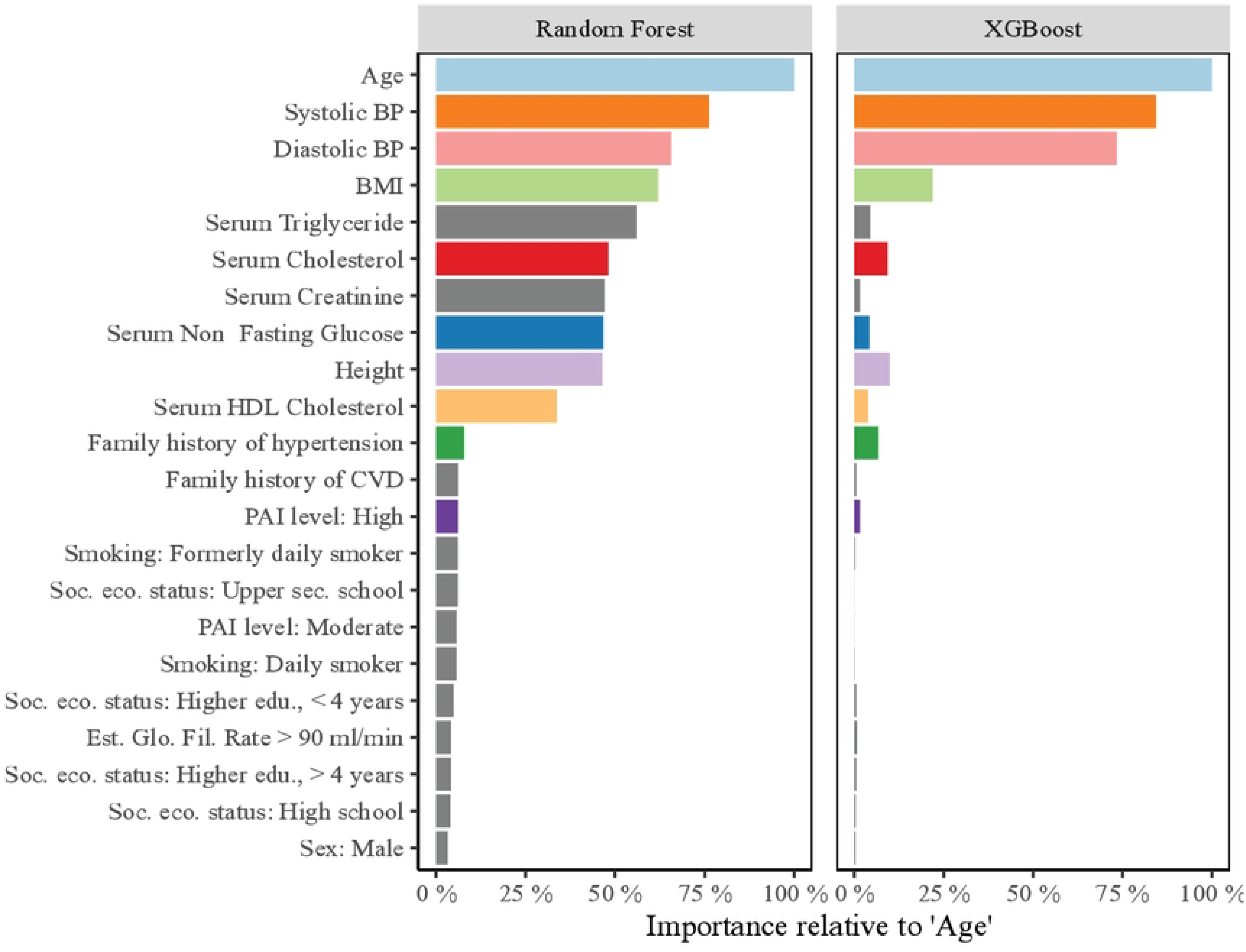
Feature-importance calculated by Random Forest and XGBoost. Permutation importance calculated by XGBoost and RF models after model fitting on the training set, sorted by importance in Random Forest. Normalized relative to ‘Age’, which was the highest ranked feature in both models. Coloring of features follow the legend of Fig A in S3 Fig.

## Discussion

In this study we have performed a thorough comparison of multiple machine learning algorithms for creating risk models for hypertension incidence. With a selection of machine learning models with distinctive characteristics, we produced well-performing models in terms of discrimination, calibration, and net benefit. This study marks the first attempt at applying machine learning methods and evaluating their performance on discrimination, calibration, and clinical usefulness within the same study. Further, this study is the first using a Norwegian cohort for constructing any risk model for hypertension incidence. In reviewing the literature, this is only the third time a Scandinavian population has been used (25,29). As these included genetic data in their models which was not present in our study data, we were not able to externally validate the model produced using other Scandinavian cohorts.

More complex machine learning methods performed better on average on each of AUC, Brier Score, and ICI, however, the apparent benefit was small. While the XGBoost model achieved the best mean AUC and Brier Score, the difference versus the much simpler elastic regression model was small, and far smaller than the variation induced by bootstrapping on the testing set. Comparing the AUC scores we achieved in this study versus a meta-analysis of the literature, we find that it is close to the expected mean AUC for a hypertension risk model (5–7). Relative to other studies, the small differences in discrimination between the elastic regression and the best performing machine learning model, is similar to that found in other studies (11,21,23).

The HUNT Study offers a dataset with large sample size. Evaluations in cross-validation and testing data yielded similar model performance indicators. This suggests that the sample size was sufficient for our study and chosen validation scheme. However, we note that we have a longer time frame than most other risk prediction models in the literature. In addition, there was a slight majority of women versus men in our data due to men being more likely to decline participation and being lost to follow-up (30). As men were more likely to develop hypertension, this might have biased the outcome rate. At the same time, the overall outcome rate of 25% in the HUNT Study is close to the age-standardized hypertension prevalence in high-income countries at 28.5%, calculated using data from 2005-2014 (31). In summary, we are confident that these factors did not affect the *relative* performance of the models, which was the focus of our study.

As a reference and external validation, we included the performance indicators produced by the Framingham risk model (4). However, we should be careful when comparing its results to our newly developed models, even though the same data is being applied. It is expected that an external validation on an unseen population will produce worse performance than the internal validation of a newly developed model, regardless of how the internal validation is performed (26,27,32). Further, the Framingham risk model is less complex than some of the machine learning methods applied in this study. Hence, the Framingham risk model results are more appropriately interpreted as a lower benchmark for acceptable performance for new risk models. For a fair comparison of the Framingham risk models and the newly developed model, they need to be applied to the same data in an external validation (26).

Performance-wise, it is notable that the Framingham model’s average AUC was comparable to some of the models developed in this study. The worse indicator for calibration was more as expected (27,32). Volzke et al also found the Framingham risk model to have an acceptable discrimination, but poor calibration on their Danish cohort (25). As for clinical usefulness, the net benefit was positive, but lower than the best performing models when the risk threshold was lower than 50%. There is a negative net benefit for risk thresholds over 50% using the Framingham risk model, which is probably related to the increasing degree of miscalibration seen in its calibration plot in S11 Fig. However, the Net Benefit of the newly developed models were small, null, or slightly negative in the same regions, meaning no model was particularly useful when a threshold above 50% was applied. Lastly, we note that the Framingham risk model was developed for a shorter follow-up and that there are some differences in the study cohort, see Table in S5 Table. Most notable, despite the lower mean blood pressure at baseline and shorter time to follow-up, the outcome ratio in the Framingham risk model cohort was far higher, 45% vs. 25% in our study data. This might explain why the Framingham risk model overestimates risk in our external validation. The rate of hypertension in close family was also different, which we suspect is related to the difference in how the feature was recorded. For the Framingham risk model, parental hypertension was recorded from a separate cohort study, where the parents themselves were assessed for hypertension multiple times over a longer period (4). In the HUNT Study data, parental hypertension feature was recorded based on a questionnaire where individuals reported for their families, which might provide less accurate data. Lastly, we did make some amendments to the model itself to match our available data, see Table in S4 Table. This may have had an impact on our external validation of the model.

Model updating or recalibration may be a method for obtaining a well-performing risk model with little effort using an external model. In applying a simple recalibration using the linear predictor of the Framingham risk model we obtained a risk model with excellent calibration, see Fig G in S12 Fig. While discrimination is unaffected by our recalibration method, the recalibrated model was on average better calibrated than the new developed models (27,33). Although it discriminated slightly worse than other models, the improved calibration was the likely cause of the Net Benefit becoming on par with the best performing models in our study. Recalibrating the Framingham risk model seems to be a worthwhile alternative to develop a new risk model from scratch using the HUNT Study data.

In the sensitivity analysis, the LASSO regression model performed well on discrimination and calibration on the testing set, even as many coefficients were zeroed out. Despite the statistically significant differences between individuals whose blood pressure remained within the range of normotension and individuals who developed hypertension at baseline, the predictive power may be low for some features. The LASSO regression model with all features except linear effects of blood pressure, age, and BMI eliminated performed well when applied on the testing set, suggesting these as important features. A possible explanation to the high performance with low features might be that the zeroed-out features have non-linear effects not captured by the LASSO regression, as opposed to a small or no linear effect.

However, we saw the same features emphasized as the most important in the XGBoost and RF models. Both methods are capable of learning non-linear effects from their inputs. The RF importance did emphasize more features than those mentioned, e.g., all serum biomarkers, and we note that these constitute all the numerical features used in this study. However, the testing set performance of the RF model was not consistently better than the other models. On average, the RF model had better calibration performance but with poorer performance on discrimination. Hence, the serum biomarkers may be regarded as important by the RF model without having much predictive power.

There are several limitations of our study. We had available 17 diverse features, including biomarkers and other easily obtainable clinical features from the HUNT Study data. The inclusion of other types of features such as genetic information, more comprehensive socioeconomic information, or information on diet could have made a difference on model performance. This became clear in the sensitivity analysis, in which only a small subset of features was determined as important. For example, Niu et al showed that the addition of a genetic risk score improved the performance of their machine learning models, but not the less complex Cox regression model (24). However, in a recent meta-analysis, the addition of genetic information was found to not improve discrimination of the included risk models (7). The latter models were largely developed using established statistical methods which included linear effects of the genetic information. Hence, which features should be included to improve predictions of incident hypertension remain unclear and a topic of further study. The second limitation was our imputation procedure: We did not perform *multiple* imputations to capture uncertainty in the imputation method. This is suggested by guidelines for developing risk models, but we refrained from doing so due to high computational costs (32). However, all models were subjected to a rigorous cross-validation scheme during development and bootstrapping of their performance on the testing set. Further, the missingness was low, with only ‘Family history of hypertension’ having a missing rate above 4%. While some variation in our imputation scheme may not have been captured, we do not think it would impact the performance of the developed models relative to each other. Third, our study is restricted to comparing the relative performance of various modelling methods for the HUNT Study data. Other relevant factors for the practical use of machine learning models are the applicability and transparency of the model (34). While many risk models are developed using relatively simple methods like logistic regression, user-friendly risk score sheets are often provided for simple application of the model for health practitioners (32). As a contrast, a computer client would be needed to calculate predictions using the XGBoost, RF, SVM, and the KNN models (35). In addition, exactly how XGBoost, RF, and SVM work for individual predictions is often complex and difficult to discern. While some argue that auxiliary methods may be helpful, other suggest avoiding use of “black-box” model predictions for high-stakes decisions (36,37). However, for these considerations to be of interest, the validation performance using more complex models should be superior to other models to justify their use. In this study, the machine learning models did not outperform models developed using less-complex methods. Lastly, we also note some limitations in the interpretation of decision curves, and how they relate to clinical usefulness. We refer to Kerr et al. and Vickers et al. for details [39], [40].

The main strength of our study is that we have taken great care in ensuring that all aspects of model-fitting and evaluation were as similar as possible for all modelling methods. This ensures that any variation seen in results or feature importance between methods are due to the characteristics of the modelling methods themselves. Further, a large sample size ensures that our fitted models were robust, with similar performance in both cross-validation and testing. Furthermore, we evaluated our models with respect to discrimination, calibration, and clinical usefulness. The similarity in our scheme for model-fitting and evaluation allows us to feel confident about comparing the performance of the developed models relative to each other. Lastly, we evaluated the externally developed Framingham risk model as an alternative to developing a new risk model from scratch. However, for application of the developed models as risk prediction models and to assess their generalization to new data, a separate validation on new unseen data would be necessary (26,27).

In conclusion, we have developed and compared the performance of hypertension risk models using different machine learning methods. While more complex methods displayed good discrimination and calibration, they did not consistently outperform a logistic regression model fitted with elastic regularization. We found the externally developed Framingham risk model to produce almost as good discrimination scores as the newly developed models. The original model overestimated hypertension risk in the HUNT Study, but this was amended by a simple recalibration to our data. In our sensitivity analysis, the features age, systolic blood pressure, diastolic blood pressure, and BMI was found to be particularly important compared to the other included features.

## Materials and methods

### Data

A dataset was derived from The Trøndelag Health (HUNT) Study, based in the now former county of Nord-Trøndelag in Norway. The HUNT Study constitutes a large population database for medical and health-related research based in four health surveys over four decades (38). Specifically, baseline data was collected from HUNT2 (1995-1997) with endpoint derived from the follow-up in HUNT3 (2006-2008).

We included individuals (>20 years of age) participating in both surveys:

- With complete information on blood pressure measurements and use of blood pressure medication at baseline and follow-up,
- without missing information on diabetes or history of cardiovascular disease (CVD) at baseline,
- with a blood pressure below the hypertension threshold and being free of blood pressure medication, CVD, and diabetes at baseline.

Blood pressure measurements in the HUNT Study were performed three times per survey, with the initial measurement used to calibrate the measurement device (38). The recorded pressure was the average of recording two and three. Hypertension status was determined following the ESC/ESH guidelines, i.e., a systolic pressure above 140, diastolic pressure above 90, or usage of blood pressure medication (39). The process of applying exclusion criteria and dataflow is shown in S10 Fig. In total, 23 722 individuals were found eligible for this study. The features available for our study are well-established risk factors of hypertension and CVD and commonly used in risk modelling of incident hypertension (7,39). We estimated physical activity by a novel physical activity metric, Personal Activity Intelligence (PAI). The PAI algorithm converts self-reported leisure time physical activity to an average weekly PAI score for the last year (40–43). The HUNT Study protocol have been described in detail by Åsvold et al. and more information about how features were collected can be found in Table in S1 Table and at https://hunt-db.medisin.ntnu.no/hunt-db/#/ (38). All participants provided informed written informed consent. This study was approved by the Regional Committee on Medical and Health Research Ethics of Norway (REK; 22902; 2018/1824). Data can be obtained upon approval from REK and HUNT Research Centre. For more information see: www.ntnu.edu/hunt/data.

The eligible cohort was stratified on outcome status (normotension; hypertension) and described by summary statistics and missing rate in Table in S2 Table. We applied unpaired t-tests or chi-square tests as appropriate to detail significant differences between those whose blood pressure remained within range of normotension or developed hypertension.

### Model development

To minimize the risk of providing overoptimistic results, we used a thorough development and validation scheme. First, we divided the available dataset randomly into a training and testing set by a 7:3 ratio. We applied unpaired t-test and the chi-square test for evaluating differences between the training and test set. Second, we applied a 4-fold cross-validation scheme on the training set to select hyperparameters for our modelling methods, shown in Fig 5. The combination of hyperparameters that produced the best mean test fold performance during cross-validation was selected for each method separately. Lastly, to produce the final model for each method, the model was fitted using the whole training set with the selected hyperparameters, see Fig 6.

**Fig 5.**
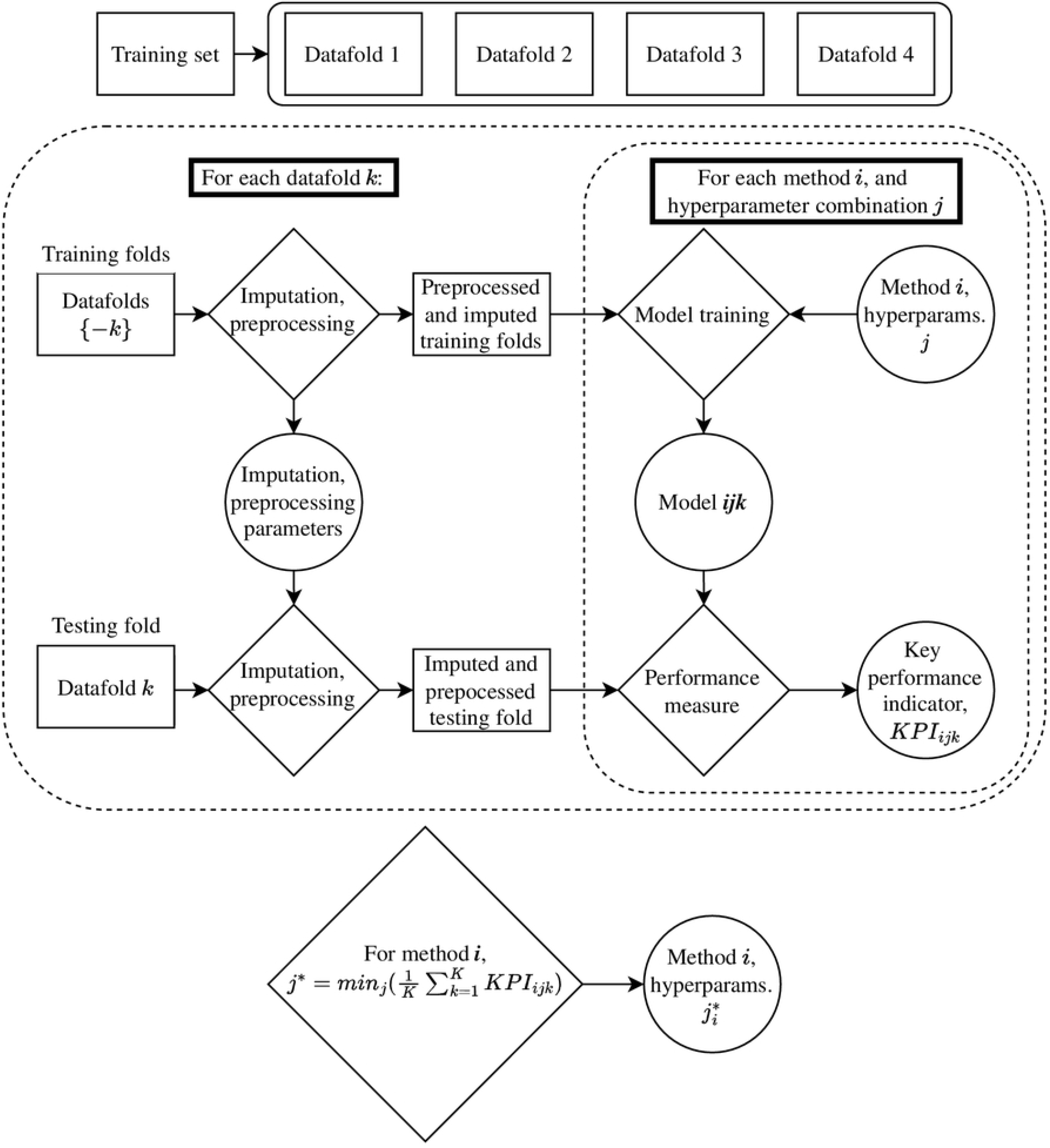
Cross-validation scheme for selecting hyperparameters. The Key Performance Indicator (KPI) used to select hyperparameters was the mean out-of-fold Brier Score.

**Fig 6.**
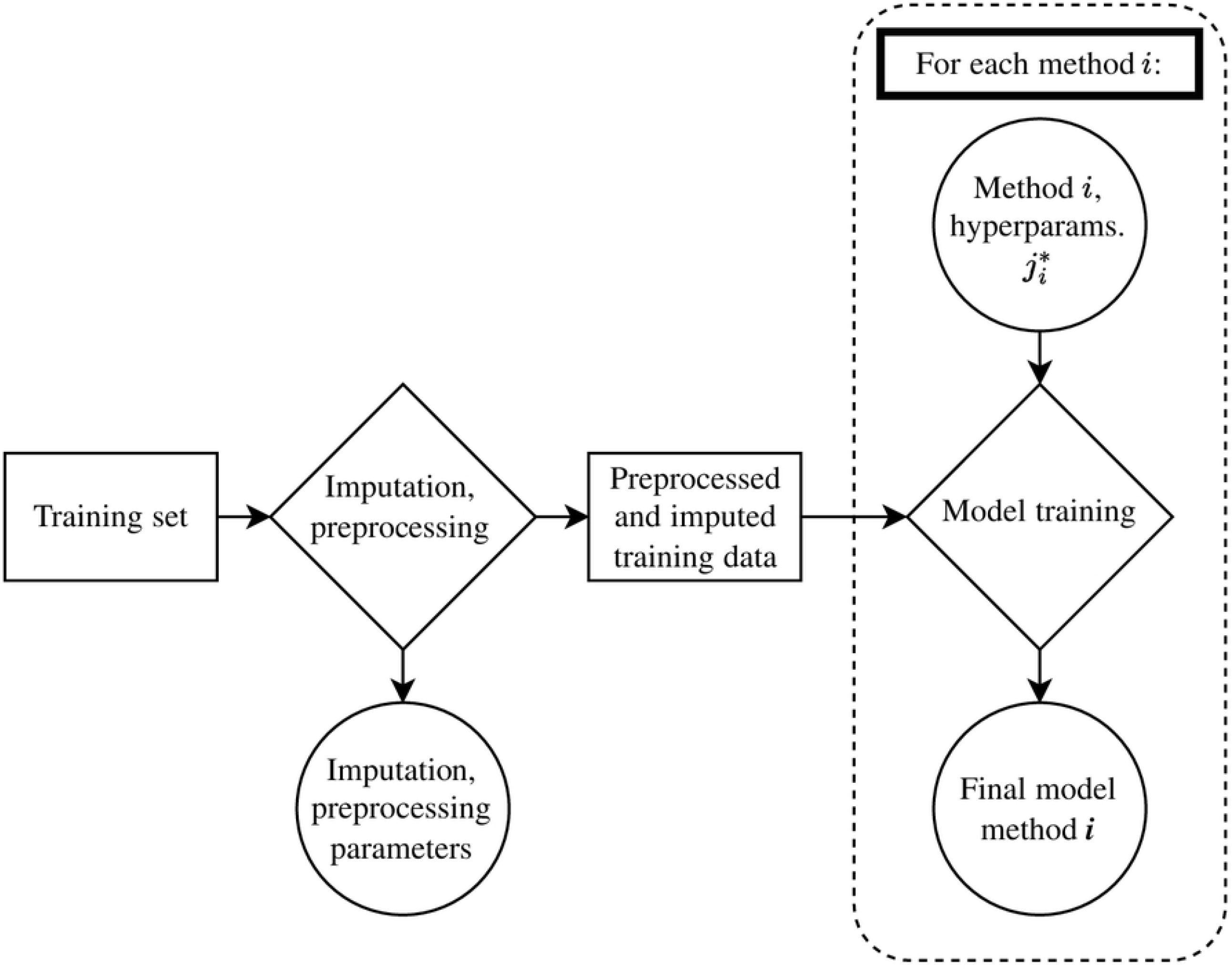
Model-fitting and learning of imputation and preprocessing parameters. Each method was fitted using the optimal hyperparameters selected in cross-validation.

### Model validation

The final models were applied on the testing set to evaluate their performance, see Fig 7. To account for variations in the data, we used a simple bootstrap scheme: The testing set was resampled with replacement 100 times, measuring the performance of each model on each resampled testing set. For each final model, the performance indicators were summarized by its mean and standard deviation from the 100 evaluations.

**Fig 7.**
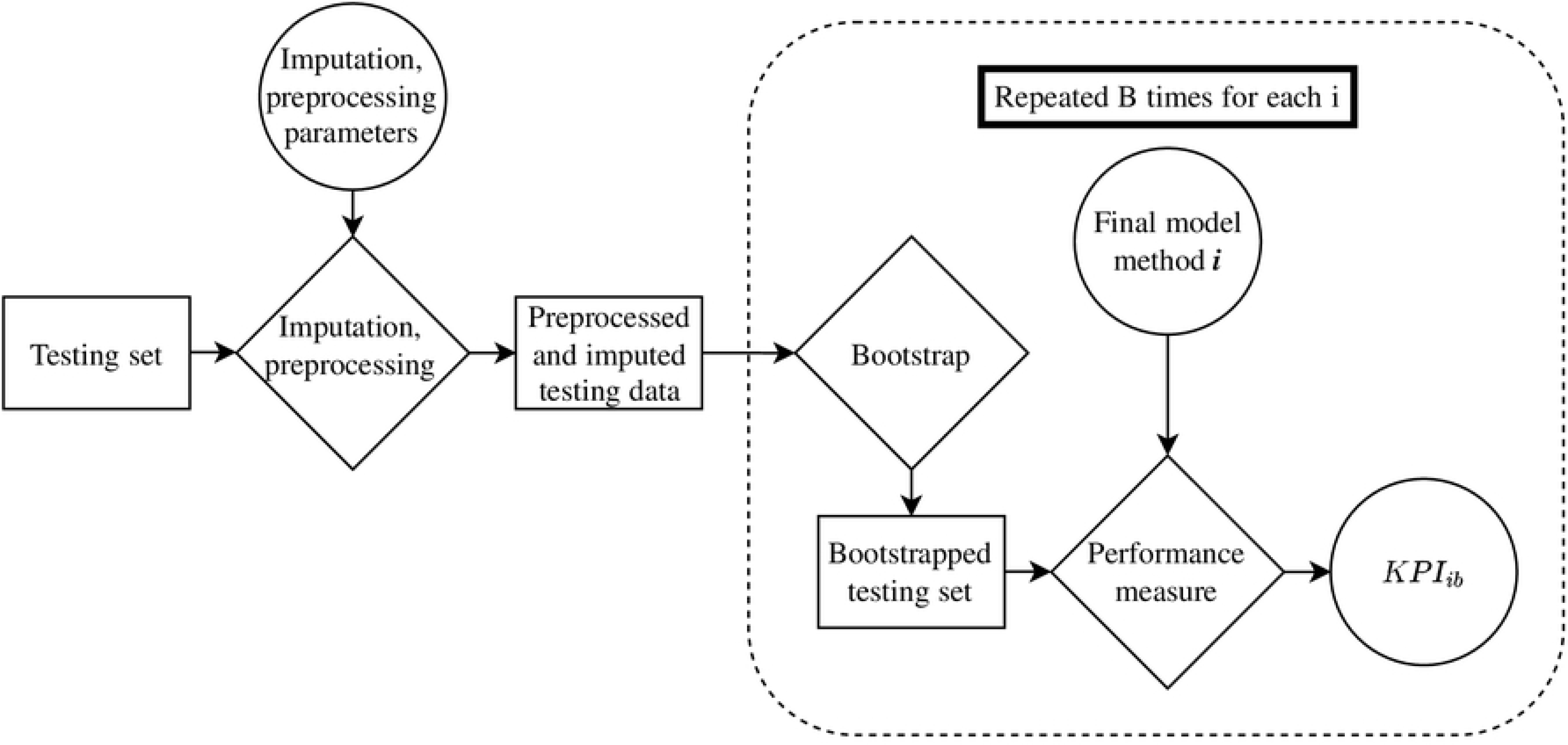
Bootstrap scheme for evaluating performance on the test set.

To ensure equal conditions for our modelling methods, we used the same data folds in cross-validation of all methods and measured model performance on the same bootstrapped testing sets. Any variation induced from the random sampling of data is therefore equal for all modelling methods, which allows for an accurate comparison of model results.

### Preprocessing and missing data

As part of the model development and evaluation, the data was preprocessed by standardizing the numerical features and one-hot-encoding the categorical or nominal features. In the case of missing feature values, we used a bagged decision tree model to impute the missing values (44). The imputation model was blinded from the outcome and learned to predict the missing feature using the other features in the dataset. Both preprocessing and imputation requires determining some parameters: For standardization, the mean and standard deviation on each feature need to be determined. For imputation, the bagged decision trees models must be fitted. To avoid ‘data-leakage’, these parameters were learned without using data the models were evaluated on. In short, the parameters and imputation method were determined in the cross-validation scheme using only the training data folds before being applied on the testing fold. Likewise, when fitting the final models, the standardization parameters and imputation model was learned using only the training set.

### Modelling methods

We included several popular and frequently used machine learning algorithms as modelling methods. In addition, we included the logistic regression method, with and without elastic regularization. The learning algorithms used were the logistic regression with and without elastic regularization (referred to as elastic regression), eXtreme Gradient Boosting algorithm (XGBoost), regularized Random Forest (RF), Support Vector Machine (SVM), and K-Nearest Neighbor (KNN) (44–48). Notably, we did not include any machine learning method using neural networks algorithms, as a recent review suggest that they display less than optimal performance on tabular data, which we employ in this study (49). Features were included in the models without defining any interactions, for simplicity.

Depending on the algorithm, the different methods included necessitated the selection of multiple hyperparameters. For each hyperparameter, a sensible range of possible values was defined, and search strategies was employed to select a value from these ranges. For the XGBoost, RF, and SVM modelling methods we employed a sampling of these ranges. In total, 70, 30, and 30 combinations of hyperparameters were sampled for the XGBoost, RF, and SVM methods, respectively, and used as candidates in the cross-validation scheme. For elastic regression and KNN, we employed a grid search where all combinations were trialed in cross-validation. The rationale behind different sampling strategies was motivated by the higher training time and computational cost of XGBoost, RF, and SVM methods compared to elastic regression and KNN. The hyperparameters, ranges and search strategies are described in Table in S3 Table.

### External models

We searched the literature for existing risk models allowing validation: Using similar features to those available in the HUNT Study data; reporting model performance; and suitable for the 11-year follow-up period between baseline and outcome. From this, we found four articles detailing such models: One being the Framingham risk model, and the three others being refitted versions of the Framingham risk model (4). We therefore included the Framingham risk model in our full model evaluation, i.e., calculated its performance on the test set along with our newly developed models. As it is expected that an external model will be poorly calibrated, we perform a simple recalibration using the linear predictor of the Framingham risk model (27,33). We learned the recalibration parameter on the training set and reported the recalibrated models performance on the test set. We did not perform any further recalibration, e.g., re-estimating coefficients. This was due to already including a simple linear predictor in our hyperparameter search for the elastic regression, i.e., elastic regression with ‘lambda’ hyperparameter set to zero, which reduce to unregularized logistic regression. We also applied the original Framingham risk model on the whole dataset, after imputation. The model, with recalibration and adaptations made for it to suit our data can be found in Table in S4 Table. We compare key aspects of the data from the Framingham risk model development study and the HUNT Study in Table in S5 Table.

### Sensitivity analysis

As sensitivity analysis, we fitted Lasso logistic regression models on the training data with increasing regularization penalty and evaluated their performance on the test set (50). We employ all features as in the model development, without interactions. With increasing penalty, less-important coefficients in the model-fitting will be shrunk towards zero. By looking at which degree of penalty the different coefficients are eliminated, we may see which features are of most importance in model-fitting for the logistic regression model. To compare, we investigated the feature importance calculated during model-fitting on the training data for the XGBoost and RF methods.

### Performance indicators

A risk models performance is primarily quantified by indicators for discrimination and calibration. In this study, we evaluated the models by the *Area Under the receiver operator Curve* (AUC). The AUC is a frequently used in literature on risk models for hypertension (7,15). We also captured the models overall performance by the Brier score, which is a proper scoring function (15,51). Further, calibration was assessed both graphically using smoothed calibration plots, and summarized using the *Integrated Calibration Index* (ICI). The ICI measures the deviation of the smoothed calibration curve of a model versus a perfect, straight, diagonal calibration line, weighted by the distribution of the model’s predictions. A low ICI score indicates that the model is well calibrated for the risk percentiles the model frequently provided predictions for in the data (52,53). Lastly, we evaluate the clinical usefulness of our models compared to sensible benchmarks graphically by presenting the *Net Benefit* plot derived from the testing set. The net benefit complements the smoothed calibration curve in assessing clinical usefulness (15,16). The benchmarks compared against in the Net Benefit plot was 1) treat all individuals as ‘Hypertensive’, 2) treat all as ‘Normotensive’, and 3) predict and treat an individual as ‘Hypertensive’ if they were prehypertensive, i.e., systolic BP above 130 or diastolic BP above 80 mmHg, at baseline, and ‘Normotensive’ otherwise.

In addition, we also reported some performance indicators frequently used in machine learning literature: The area under the Precision-Recall curve, the F1 measure, sensitivity, specificity, positive predictive value, negative predictive value, and the Matthews correlation coefficient (8). For the performance indicators where predictions needed to be either “Normotensive” or “Hypertensive”, we assigned all individual predictions below the outcome rate (24.36%) of the training data as ‘Normotensive’, and above as ‘Hypertensive’.

As a common criteria for choosing the optimal models during cross-validation, we used the Brier Score, as it is a proper scoring rule regardless of modelling method (54). Hence, the optimal set of hyperparameters during cross-validation was the one producing models with the lowest Brier Score.

### Software and reporting

We have used the RStudio IDE and R for implementing the modelling algorithms and data-processing. The following R-packages were used: *skimr* for data exploration, *randomForest, RRF, glmnet, Matrix, xgboost, plyr, kernlab, class* and *caret* for modelling algorithms, *recipes* for preprocessing, *glmnet* for sensitivity analysis, and *ggplot2, ggextra, patchwork, ggh4x* and *dcurves* for graphics (55–70).

To ensure a high degree of transparency and quality in reporting, we strived to follow the *Transparent Reporting of a multivariable prediction model for Individual Prognosis Or Diagnosis* (TRIPOD) (32). We note that a guideline is being developed for prognostic prediction model studies based on artificial intelligence, which may have been more relevant for our study (71). However, the guideline was not available at the time of writing. A form detailing adherence to the TRIPOD guidelines for the development of the new models and the validation of the Framingham risk model has been attached as Supporting Information, see Files S13 and S14.

## Data Availability

The Trøndelag Health Study (HUNT) has invited persons aged 13 - 100 years to four surveys between 1984 and 2019. Comprehensive data from more than 140,000 persons having participated at least once and biological material from 78,000 persons are collected. The data are stored in HUNT databank and biological material in HUNT biobank. HUNT Research Centre has permission from the Norwegian Data Inspectorate to store and handle these data. The key identification in the data base is the personal identification number given to all Norwegians at birth or immigration, whilst de-identified data are sent to researchers upon approval of a research protocol by the Regional Ethical Committee and HUNT Research Centre. To protect participants’ privacy, HUNT Research Centre aims to limit storage of data outside HUNT databank, and cannot deposit data in open repositories. HUNT databank has precise information on all data exported to different projects and are able to reproduce these on request. There are no restrictions regarding data export given approval of applications to HUNT Research Centre. For more information see: http://www.ntnu.edu/hunt/data

## Acknowledgments

The HUNT Study is a collaboration between HUNT Research Centre (Faculty of Medicine and Health Sciences, Norwegian University of Science and Technology, Trøndelag County Council, Central Norway Regional Health Authority, and the Norwegian Institute of Public Health. We thank the participants and management team of the HUNT Study. We also thank the staff at HUNT Cloud who aided us with tools for data storage and analysis.

## Online resources

To encourage dissemination of the risk models developed in this study, we created an online resource, https://github.com/filsch/hypertension_prediction_models_hunt_study, where multiple risk models and auxiliary functions are provided for easy utilization by external researchers. Some example data is provided to ensure the right formatting of data to be used in the models. The risk models included in the resource are the XGBoost, RF and the elastic regression model. In addition, a logistic regression model using fewer features derived from the sensitivity analysis, the adapted Framingham risk model, and the adapted Framingham risk model recalibrated on the HUNT Study data are also included.

## Funding

The author(s) received no specific funding for this work.

## Supporting information captions

**S1 Table. Variable names used to construct features, as named in HUNT databank**.

**S2 Table. Feature distributions for all data and subdivided by outcome status**.

**S3 Table. Hyperparameters, candidate values, and search strategy per modelling method**.

**S4 Table. Adaptations and full model equation of the original Framingham risk model and after recalibration to the HUNT Study data**.

**S5 Table. Study and data characteristics used in developing the Framingham risk model and in this study**.

**S6 Table. Feature and outcome distributions for all data and subdivided by training and test set**.

**S7 Table. Hyperparameters selected with out-of-fold performance in cross-validation on training set**.

**S1 Table. Auxiliary performance measures calculated on the test set**.

**S9 Table. Results from sensitivity analysis in tabular form**.

**S10 Fig. Dataflow on applying exclusion criteria**.

The flow of datapoints relative to the application of exclusion criteria on the available data from the HUNT Study.

**S11 Fig. Individual calibration curves for all models using the test set data**.

Displayed as black mean curves and red shaded 95% confidence interval, calculated using pointwise bootstrapping. Dashed line indicates a perfect calibration line. Histogram of predictions on the test set displayed on top of plot, color-coded by outcome status. Color coding of curves corresponds to legend in Fig 1 and Fig 2.

**S12 Fig. Individual net benefit curves for all models using the test set data**.

Displayed as black mean curves and red shaded 95% confidence interval, calculated using pointwise bootstrapping. Color coding of curves corresponds to legend in Fig 1 and Fig 2.

**S13 File. TRIPOD form for the development of models using the HUNT Study data**.

**S14 File. TRIPOD form for the external validation of the Framingham risk model**.

**S15 File. R scripts for analysis and plotting, and datafile for plotting figures and tabular data**.

## Notes

### Competing Interest Statement

The authors have declared no competing interest.

### Author Declarations

The Regional Ethics Commitee (REK) is responsible for approving all medical research that is subjected to the law on health research. See: https://rekportalen.no/#hjem/home

